# Conundrum of re-positives COVID-19 cases: A Systematic review of Case reports and Case series

**DOI:** 10.1101/2020.12.10.20223990

**Authors:** Arun Kumar Yadav, Subhadeep Ghosh, Sudhir Dubey

**Affiliations:** Armed Forces Medical College; ACMS

## Abstract

**Introduction:** There have been case reports and case series published for RT PCR positive COVID - 19 cases that became RT PCR negative but subsequently became RT PCR positive after a symptom free interval following a negative RT PCR test. These cases may include re-positive, reactivated and re-infection cases. Hence, the systematic review to summarize and synthesize evidence from all available case series and case reports published was undertaken.

**Methodology:** The systematic review of case series and case reports was registered with Prospero with registration number CRD42020210446. PRISMA guidelines were followed for conducting the systematic review. Studies published in English language only were considered for the Systematic Review. Inclusion criteria for studies included case reports and case series which have documented cases of positive RT-PCR after a period of improvement or negative RT PCR. Reviews, opinions and animal studies were excluded. Case reports which described clinical presentation or manifestations of COVID-19 cases were also excluded from the studies. Methodological quality was assessed using modified Murad scale.

**Results:** A total of 30 case reports/case series were included in the study, wherein a total of 219 cases were included. In re-positive cases, the age range varied from 10 months to 91 years. The pooled proportion using random effects was 12% with 95% CI from 09% to 15%. Among the re-positives, a total of 57 cases (26%) of the cases had co-morbidities. A total of 51 (23.3%) and 17 (7.8%) re-positive cases had been treated with antivirals and corticosteroids respectively. Among the symptomatic cases, the disease severity was lesser as compared to the initial episode of illness. Only a few studies have confirmed the presence of antibodies after the first episode. The few studies that had done contact tracing of re-positives did not find any positive cases among those in contact with re-positives.

**Conclusion:** This systematic review presents the review of all the case reports and case series on recurrence of COVID 19 disease. Although limited evidence has been generated due to paucity of such studies and shortcomings in the study designs of case reports and case-series, nonetheless, the evidence generated can still be used in making clinical decisions and framing policy guidelines

## Introduction

Clusters of cases of atypical pneumonia were reported from Wuhan city, China in Dec 2019 in hubei province (1). The disease was later renamed as COVID-19 and the agent was later identified as Severe Acute Respiratory Syndrome Corona Virus −2(2). WHO has declared it as public health emergency of International concern (PHEIC) on 30 Jan 20 and subsequently as a pandemic on 11 Mar 20(3).

As on 16 September 2020 29,444,198 COVID cases and 9,31,321 deaths have been reported globally (4). More than nine months into the pandemic, steady accumulation of scientific data and evidence in the context of disease dynamics, transmission, pathophysiology, diagnostic and treatment modalities, a large number of existing knowledge gaps have gradually been filled; however certain aspects of the disease pertaining to immune response to the disease (Humoral versus Cellular Immunity, persistence of acquired immunity and natural immunity to the disease) are still in a nascent stage of conception. These issues assume greater importance with reports of re-activation/ relapse of the disease creating sensational headlines and the imminent development and marketing of a plethora of vaccines on the anvil.

There have been various case reports and case series published for RT PCR positive COVID −19 cases who have become RT PCR negative and again become RT PCR positive after symptom free period or RT PCR negative test. These cases may include, re-positive, reactivated and reinfection cases. It is not known whether these cases share common characteristics or may share common characteristics which may help in identification prior to their discharge. The systematic review of the case reports and case series of the re-positive may help in better understanding of the natural history of the disease. Hence, systematic review to summarize and synthesize evidence from all the case series and case reports published was undertaken.

## Methodology

The systematic review of case series and case reports was registered with the Prospero with registration number CRD42020210446. PRISMA guidelines were followed for conducting the systematic review. A detailed literature search was done till 12 Sep 2020 for studies having reported cases of COVID-19 after a symptom-free interval. The databases that were searched included Medline through Pubmed, and Cochrane databases. The key term used were COVID-19, Severe Acute Respiratory Syndrome Corona Virus, Relapse, Reactivation and Re-infection. The detailed search for Pubmed is given in supplementary table 1. Hand searches of the references of articles was also done. Observational studies including Case Reports and Case Series which have reported COVID-19 cases after a symptom free interval were taken into the systematic review. Studies published in English language only were considered for the Systematic Review. Inclusion criteria for studies includes case reports and case series who have documented cases of positive RT-PCR after period of improvement or negative RT PCR. Review, opinions and animal studies were excluded. Case reports which described clinical presentation or manifestations of COVID-19 cases were also excluded from the studies.

**Table 1:**
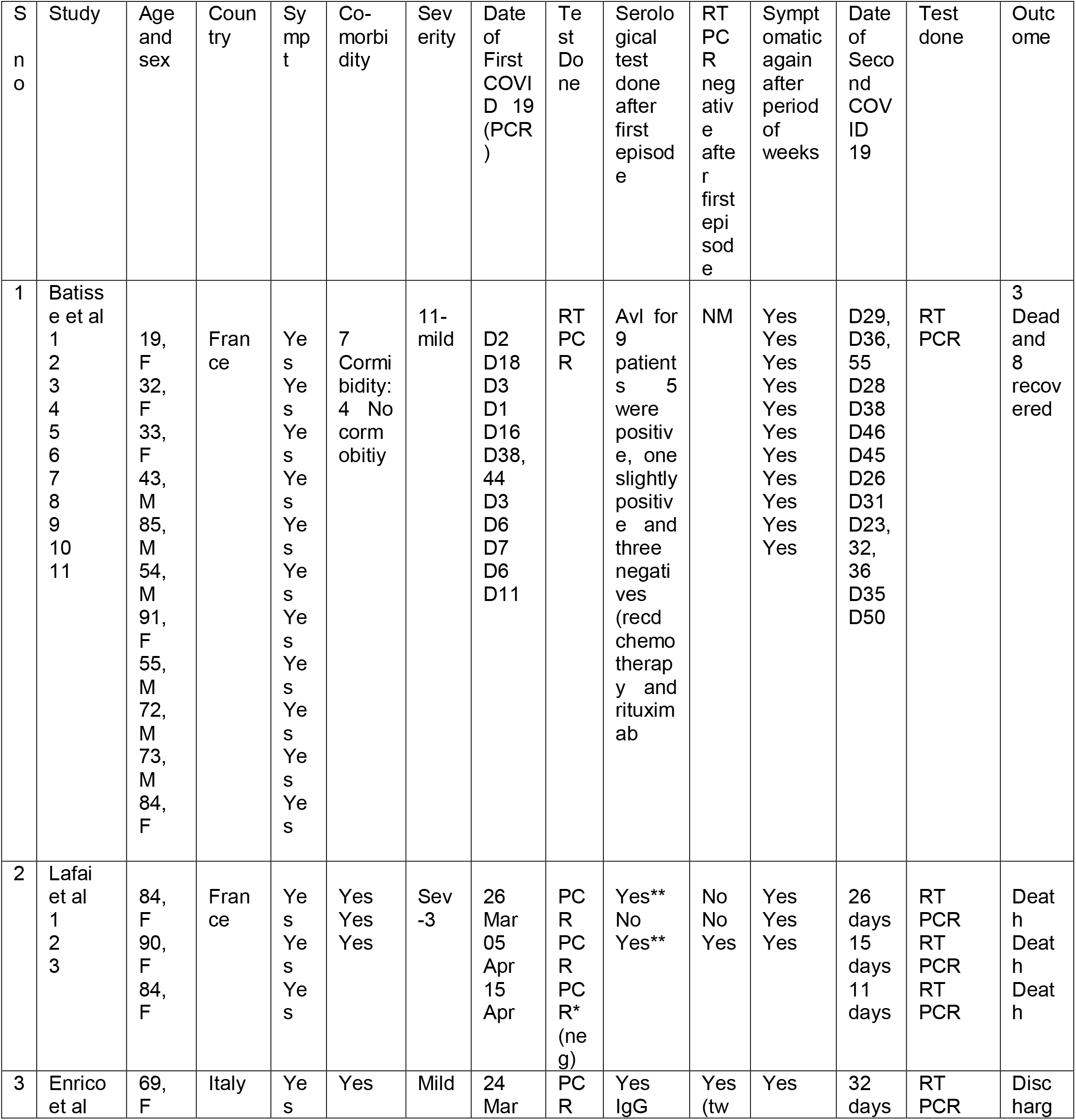
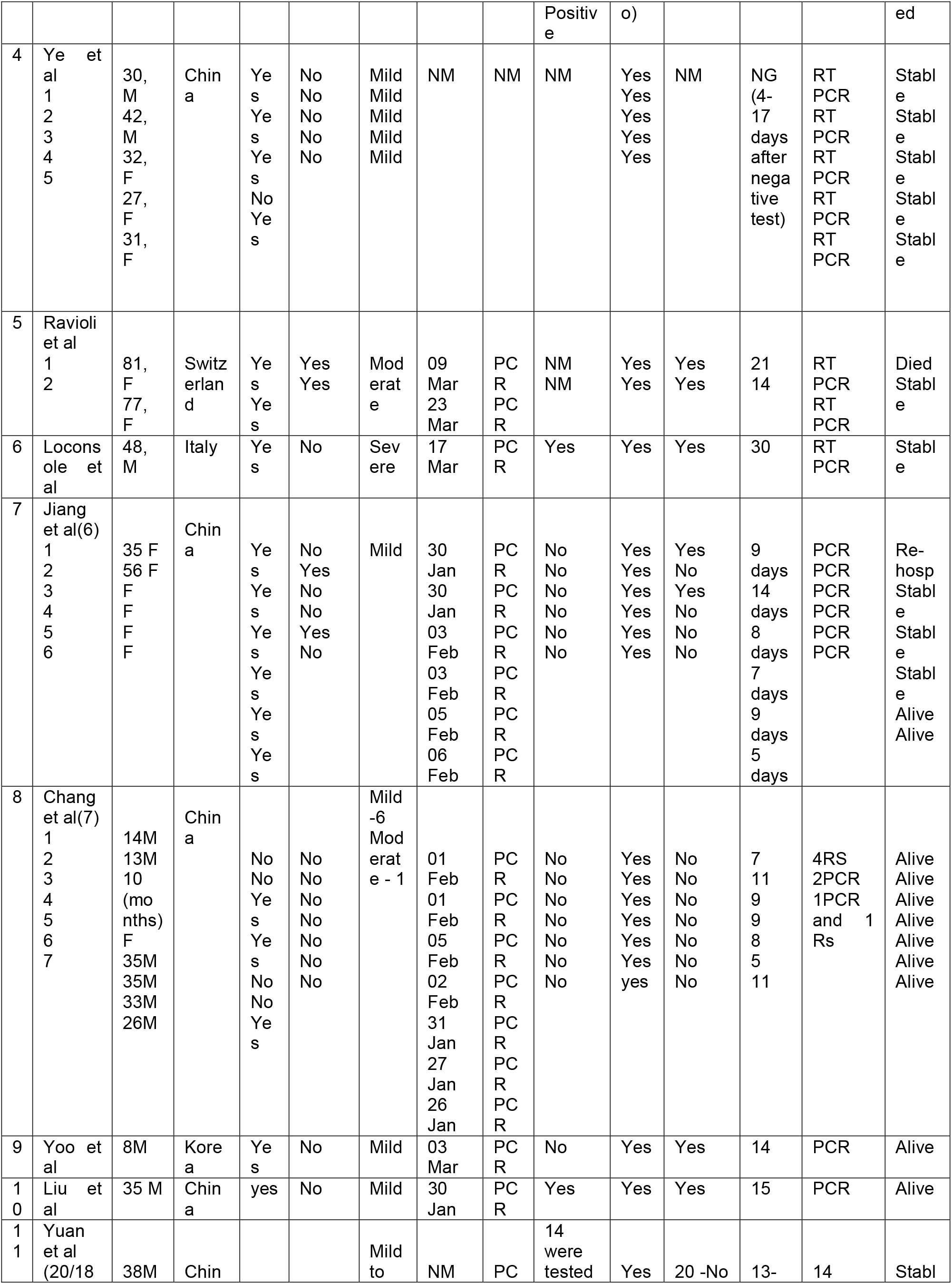
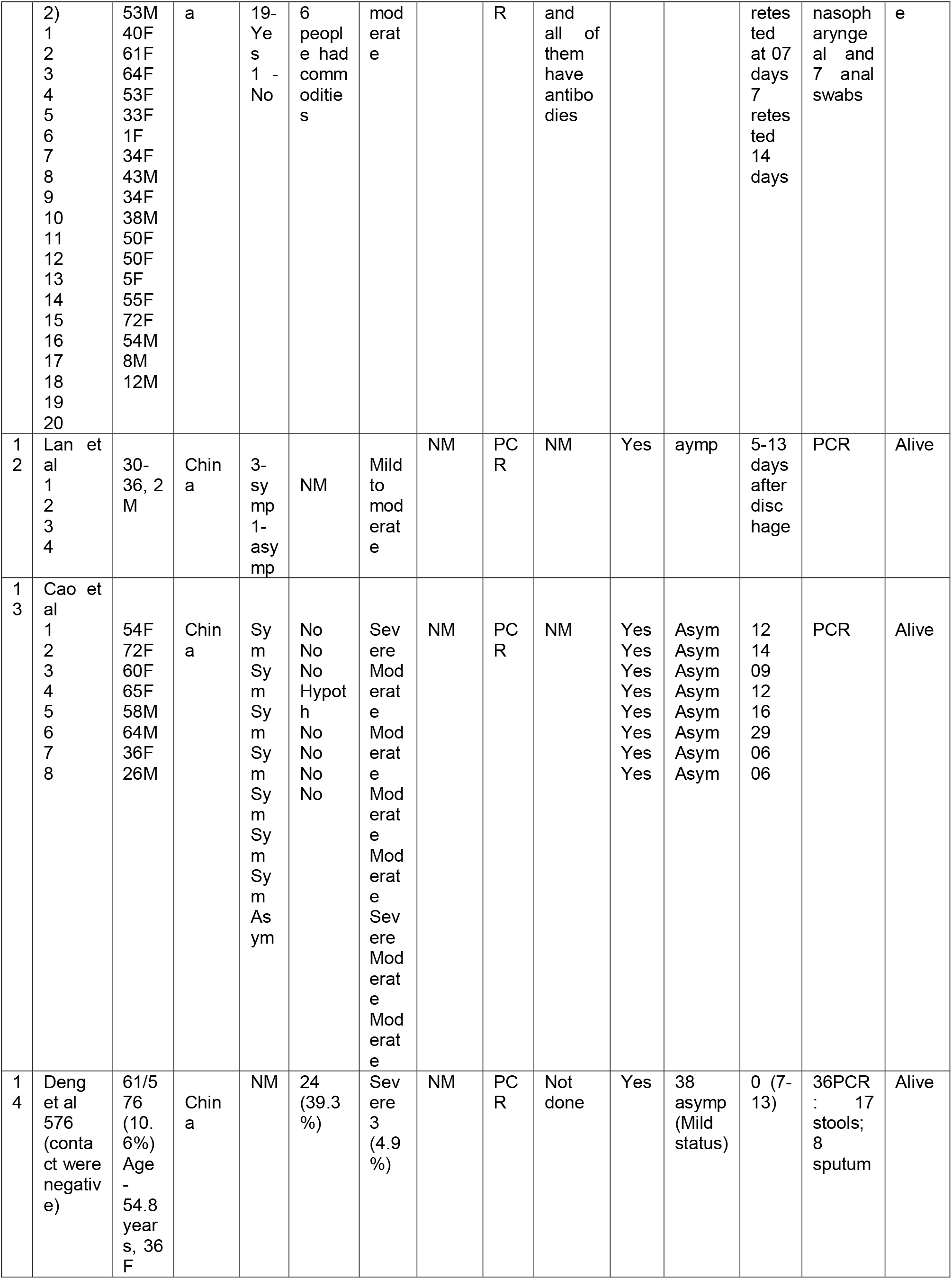
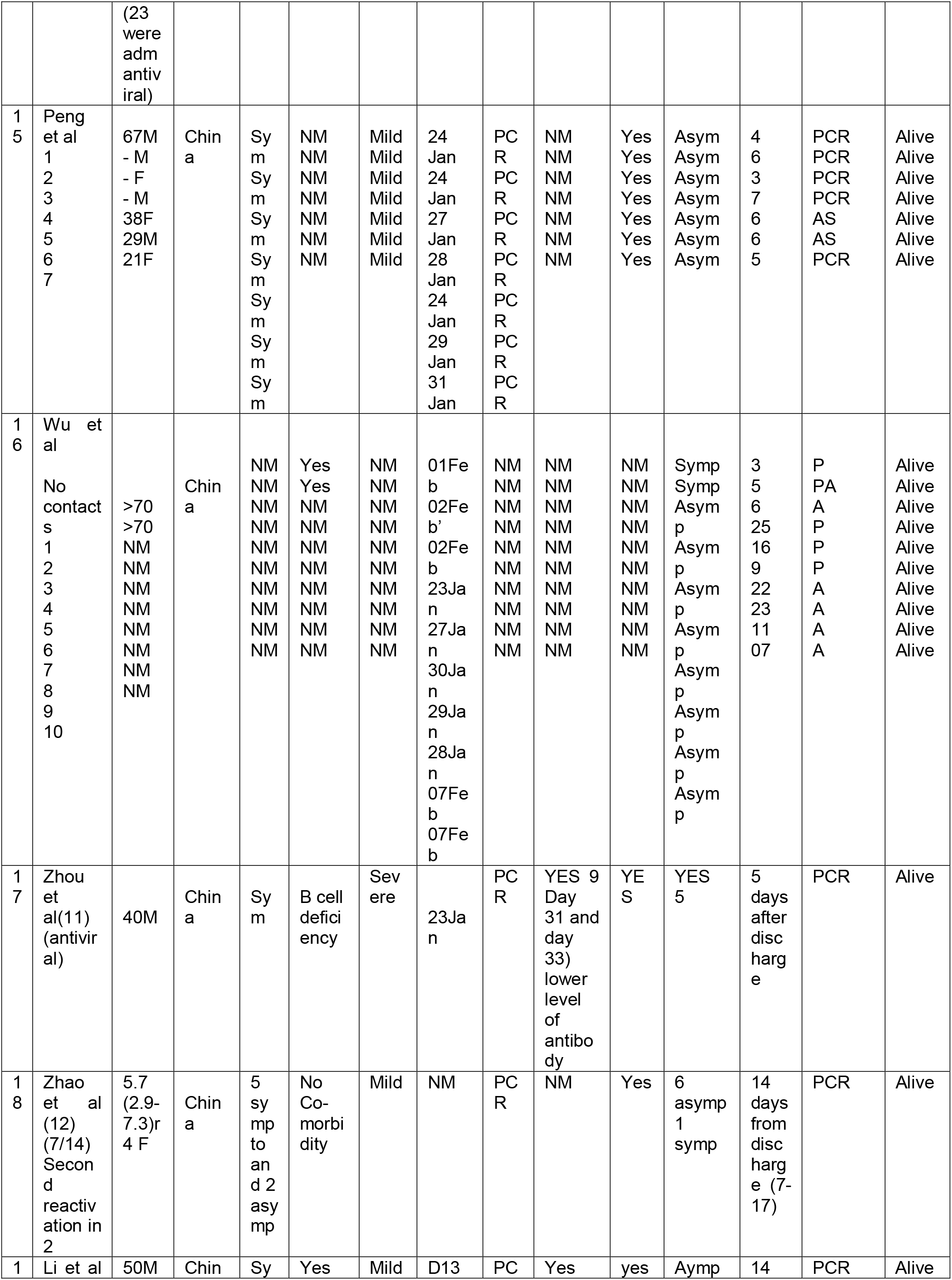
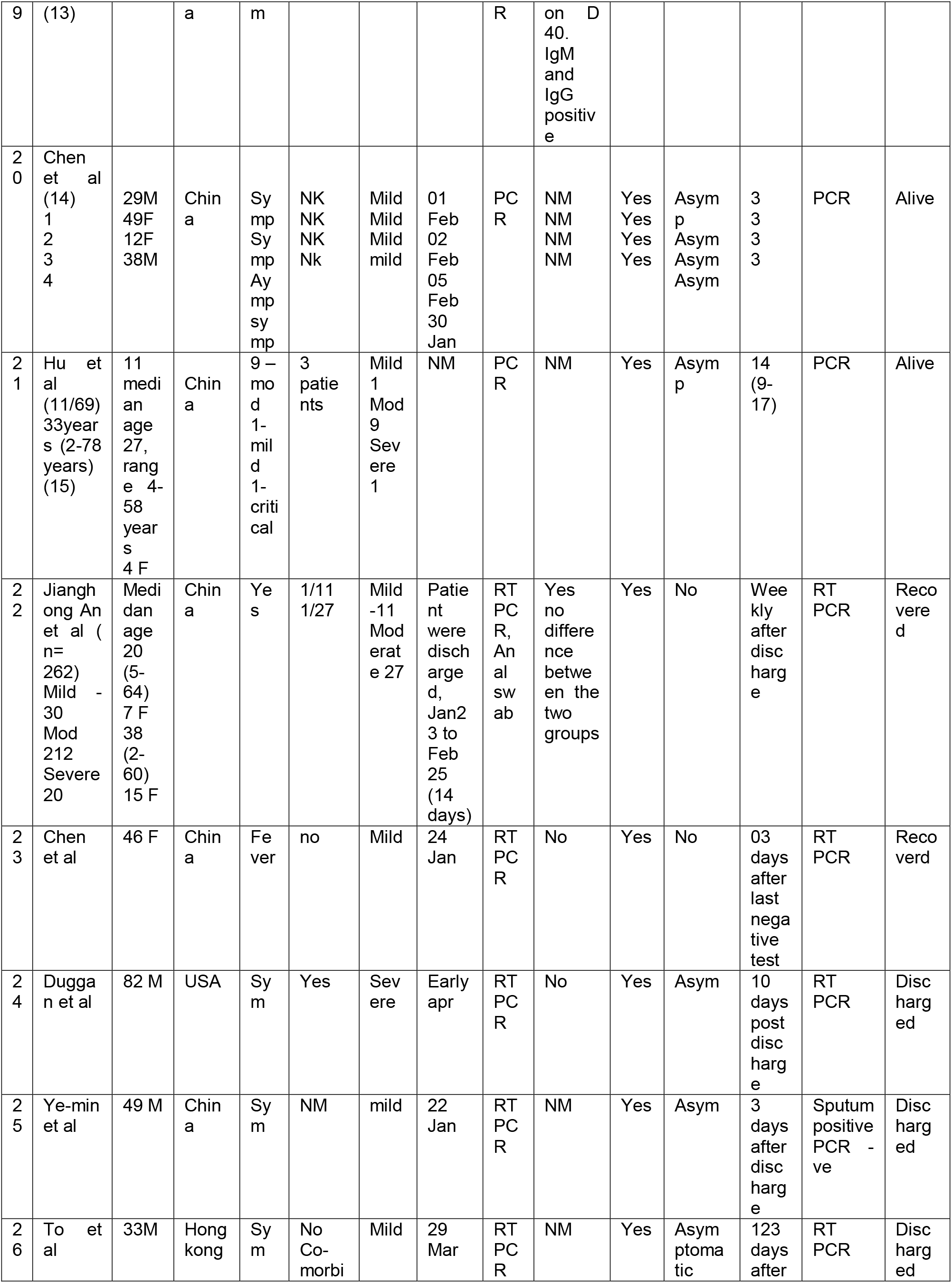
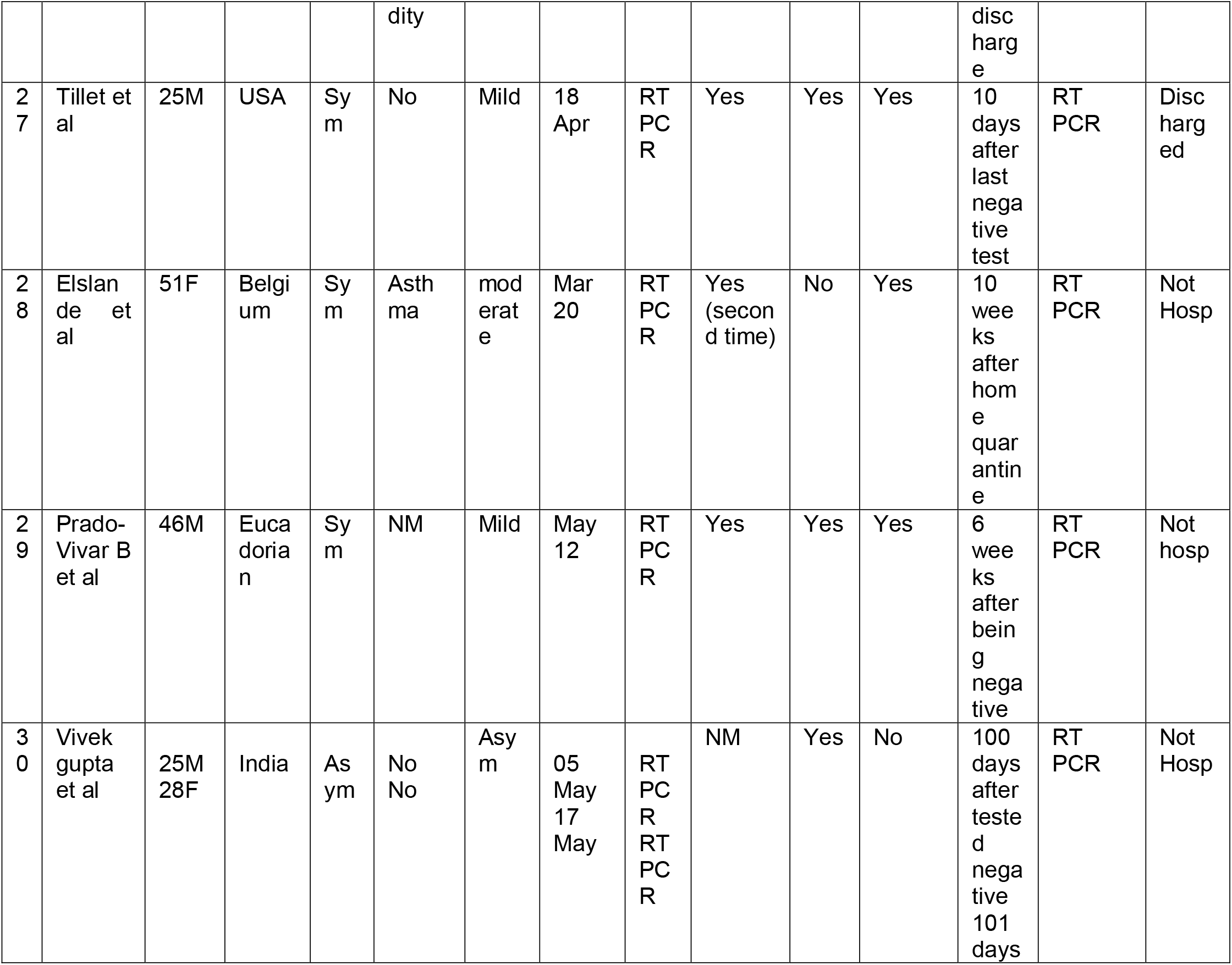
Characteristic of studies

A data extraction form was synthesized and data was extracted by two authors independently. The data items consisted of age and sex of the patients, clinical co-morbidities, date of initial positive PCR test, date of negative PCR test based on which the patient was declared as cured and date of positive PCR test in recovered individuals who reported with fresh onset of symptoms suggestive of COVID-19 infection after a disease free interval. Data on serology (If performed) and clinical outcome of patients was also collated.

Methodological quality will be assessed using the existing guidelines (5). The narrative synthesis of the results would be done. The meta-analysis technique for pooling of results would be used wherever possible.

## Results

The selection of the study is shown as PRISMA Chart in Figure 1. A total of 29 case reports/case series were included in the study. A total of 219 cases from 30 studies were included in the study. The details and characteristics of the patients in the case series and case reports are shown in table 1(6–25,25–34).

**Fig 1.**
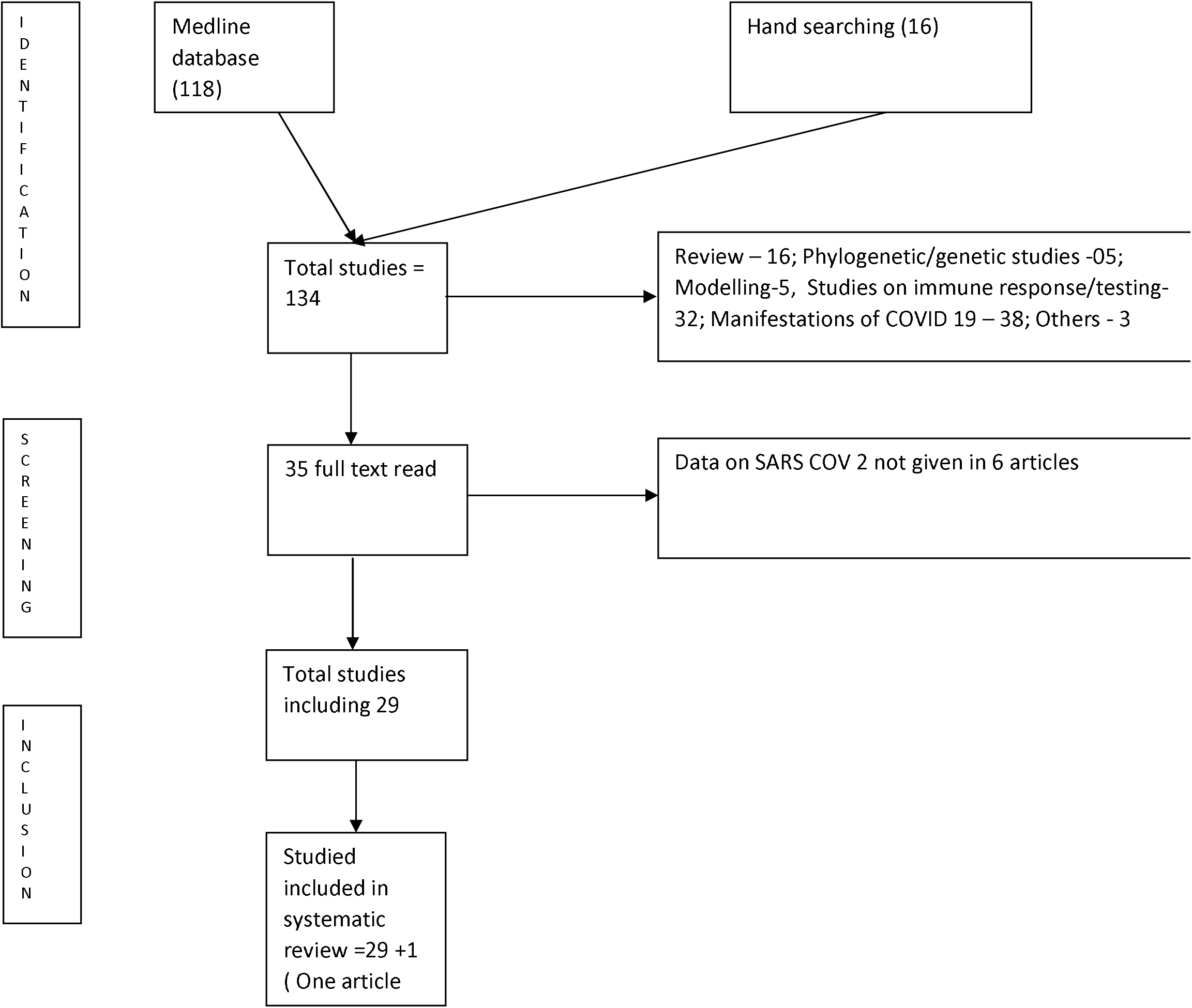
Prisma Chart for the inclusion of studies in the systematic review.

A total of eight studies gives the proportion of cases who are positive after the follow-up period which ranges from one week to seven weeks. A total of eight studies have mentioned proportion of cases who became repositive after negative RT PCR test. The summary of proportions and their pooled ratio is given in figure 2. The pooled proportion using random effects was 12% with 95% CI from 09% to 15%. All studies had follow up period of in the range 4-17 days except Jianghong et al which had followup time of 14-46 days.

**Figure 2:**
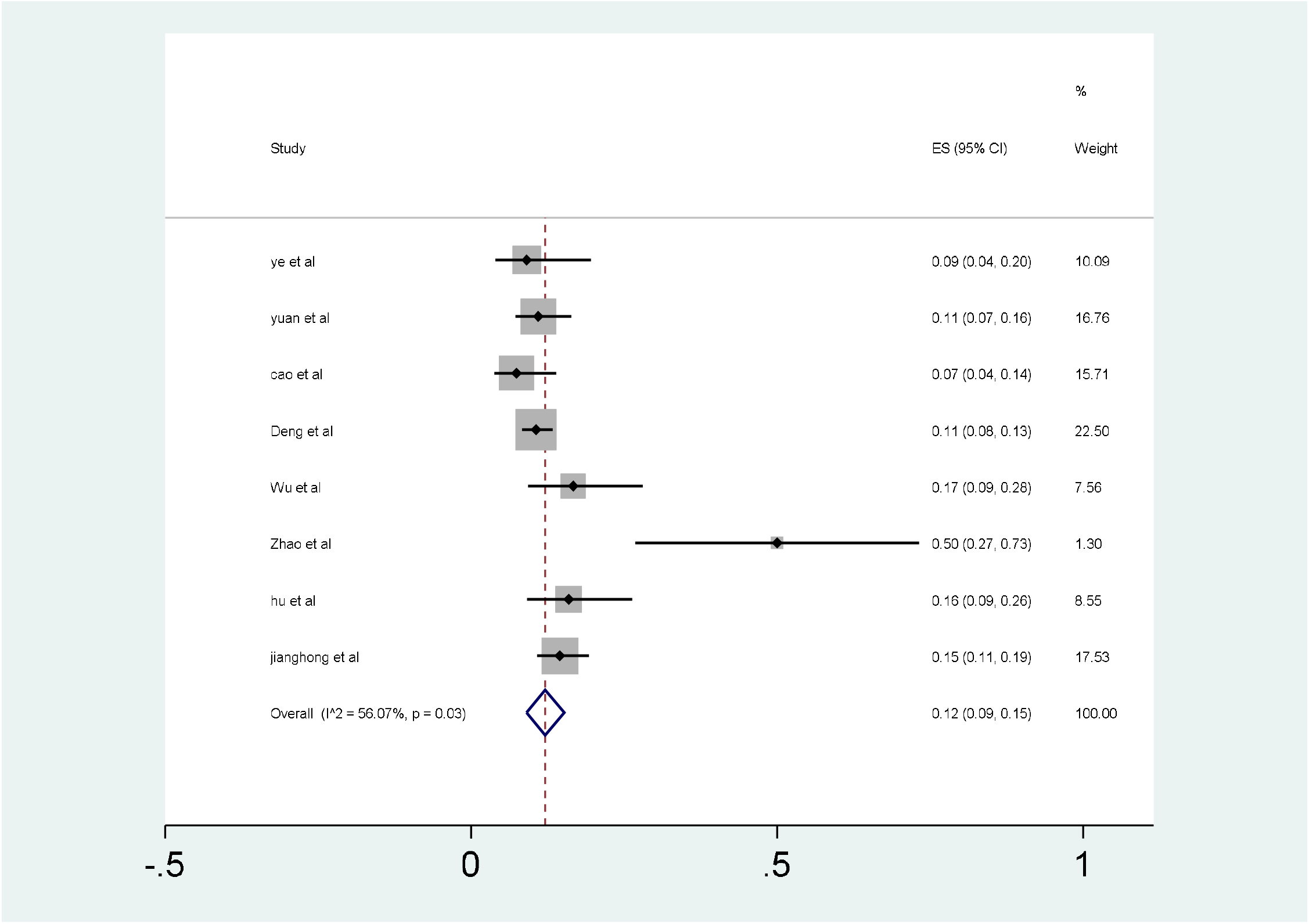
Pooled proportions from studies.

The age range of the recurrence cases varies from 10 months to 91 years of age. The pooled mean age of 195 cases was 44.3 ± 19.2 years. Zhao et al studied recurrence in children and wu et al data for combining the age was not mentioned(21,23). A total of 111 out of 209 were females. Sex was not mentioned for 10 cases.

The COVID 19 testing among discharged patients has been done from sputum (lower respiratory tract), nasopharyngeal and anal swab the details are shown in table 1.

A total of 136 (62.1;95% CI: 55.3 – 68.5)COVID Cases were symptomatic, However, the status was not known for 70 (32%) COVID cases at initial presentation. Only 12 cases (5.5 : 95% CI; 2.8 – 9,4) were of severe cases as reported in the studies. Majority of the cases (197, 89.9%) had mild-moderate presentation While the presentation was not known for 10 COVID cases. A total of 57 cases (26%) of the repositives cases have Co-morbidities. A total of 51 and 17 COVID cases had taken antivirals and corticosteroids respectively. A total of 64 (29.2%) reported COVID cases were symptomatic in second episodes, 150 (68.5%) were asymptomatic and the status of five was unknown. The range of days for positivity varied from 03 days to 101 days after discharge.

Only a few studies confirmed the antibody presence after first episode (table 1). However even after development of antiboides studies have reported re-positivity. Only a few studies have looked into genetic analysis of the SARS COV2 to confirm the reinfection.

Few studies had done contact tracing of repositives, however to date did not find any positive cases in contact with repositives.

The mortality has been reported in seven repositives cases. The age range of these cases varies from 73-91 years. All of them had multiple co-morbidities.

A total of only 63 reported cases were symptomatic after the first episode, however, the majority of them were less severe than the first episode of COVID 19 cases. Only few studies have shown antibody formation after first episode.

The quality of studies assessed by using the modified Murad et al scale is shown in figure 3. In most of the methods selection methods are not clear and also there were no precautions taken for ruling out false positive or rule out the pathogen.

**Figure 3:**
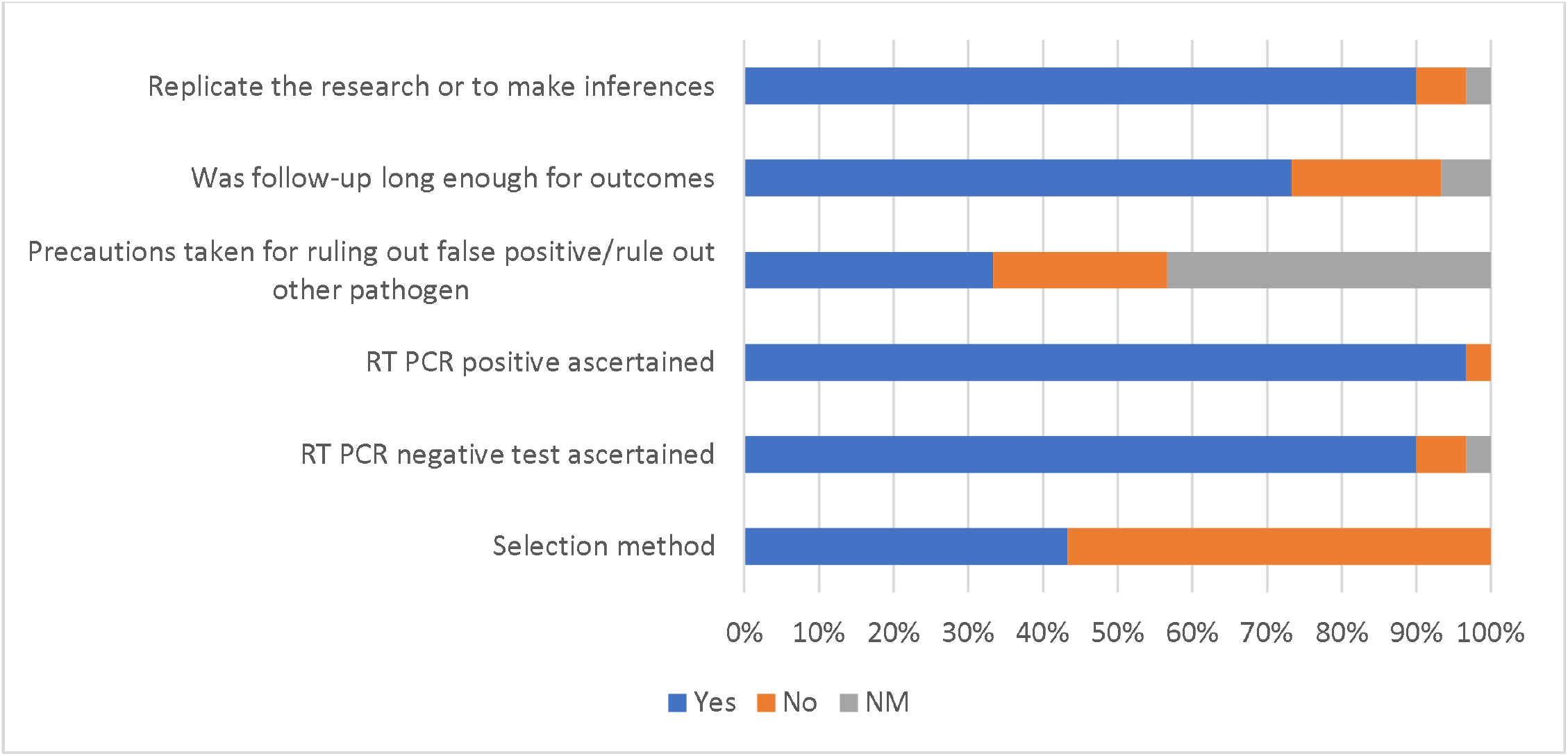
Quality of study as assessed using modified.

Korea Centers for Disease Control and Prevention reported 141 cases positive by RT PCR after they recovered from COVID-19(35). However, the probable reason given was relapse or inconsistent tests. The details were not available on the site.

Risk of bias. Though there are no set guidelines for the estimation of the risk of bias, the author feels that initial PCR positive, subsequent PCR negative, serological testing and PCR positive after symptom-free period are essential for draw conclusion about relapse or reinfection. Only case reports by Lafai et al and Enrico et al have reported negative PCR(7,8). Case reports by Batissee et al and Ravioli et al did not mention a negative PCR test after first COVID infection(6,10). Loncosole et al provided all the requiste information(11)

## Discussion

### Definition of relapse

The systematic review was done of all case reports and case series to identify common characteristics and evidence available for repositives of the cases. Though in literature search we found evidence of repositives of COVID 19 cases after symptoms free and negative RT PCR test, yet it is difficult to ascertain whether it was due to continuous shedding of the virus, relapse, or reinfection of the virus. Only six studies are there which have done the genetic analysis of the COVID 19 virus and found different genetic disease among those who have recovered from the COVID-19.

The recurrence has been observed across all ages from 10 months to 91 years of age. The mortality is seen in older cases with multiple co-morbidities in consonance with the primary infection. Immunity of the individual may also influence recurrences. Hence immunosenescent of the old age as well immunosuppressants drugs may affect the recurrence. However, majority of the COVID 19 repositves cases were not given corticosteroids. Many repositives cases were also given antiviral drug. Howerver there is a limitation of making inference from the systematic review as no valid control group were not present and secondly the denominatior in case reports or case series is difficult to ascertain. The effect of other immunomodulator and antiviral drug on recurrence may be studied in well designed study.

The pooled proportion of studies that have given the proportion of COVID 19 repositives was done. Around 12% of discharged COVID 19 cases after comes out positive. The reasons may be related to Intermittent shedding of virus, the persistence of the virus, testing technique including sampling, or host characteristics. The persistence of virus in body is known phenomenon for SARS-associated Coronavirus(36). As of now there was no evidence of secondary cases from these repositives, however, the possibility of spread of infection does exist. This underlines the importance of surveillance of discharge cases of COVID 19.

Different site for sample may also have some effect as in many cases even if the sample from nasopharyngeal are negative, the samples from sputum (lower respiratory tract) and anal swab have been positive. There is an evidence that virus may be shed longer from extraphrangeal site. There are reports that of virus shedding from asymptomatic patients may continue from extrapulmonary sites in various bodily fluids (Saliva, tears, feaces, throat, or nasal discharge) for longer duration of time(37,38). Its role in re-infection is still not known. However there are few studies which suggests no role of shedding of virus on reinfection(39)

Yuan et al noted no difference between among repositves and negatives for antibody formation and also reported that aymptomatic cases may be repositives. The animal studies however have shown the prevention of reinfection with antibody formation(40) The role of antibody in virus shedding and reinfection may needs to be elucidated in further studies. Re-infection may lead to the selection of escape mutants and subsequent dissemination to the population.

Antibody dependent enhancement is a known phenomenon in viral disease, responsible for increased severity of subsequent infections, However, in this systematic review we found that majority of re-positives cases were milder than first infection. This may be due to reasons that most of the cases are not reinfection but persistence of same infection or interaction between virus-virus. A modelling for reinfection has concluded that the rate of reinfection by the recovered population will decline to zero overtime as the virus is cleared clinically from the system of the recovered class(41)

This systematic review presents the review of all the case reports and case series on recurrence of the diseases. The evidence generated may not be of high level but can still be used in making clinical decision and policy making. Clinician may suspect the COVID cases among recovered cases in which all other diagnosis has been ruled out and policy makers and public health specialist may need to modify the policy as per newer evidence. The infectiousness of these recurrence cases may need to be further explored as this will have major implication on public health policy.

Since these patients of recurrence may represent special subset of COVID cases, the findings may not be generalizable to all COVID cases. More research is needed to delineate the factors responsible for recurrence in cases. As the pandemic progress the evidence would need further revision. Nevertheless, there a strong case for proper documentation of all the cases to further refute or confirm the findings.

## Data Availability

All the data may be provided on request

